# Incidence and Relative Risk of infection with SARS-CoV-2 virus in European Soccer Players

**DOI:** 10.1101/2021.01.19.21250085

**Authors:** Thomas B. Andersen, Andreas B. Dahl, Jeppe B. Carstensen

## Abstract

The purpose of the present study was to investigate the incidence and relative risk of infection Covid-virus in soccer players. Data from five leagues was used and compared to data from the normal population in each country. Our results revealed that the relative risk was higher in soccer players in three countries when correcting for the estimated true number of infected people in the populations. We discuss that the reason for the higher incidence in soccer players is caused by the virus entering a group of players that work closely together.

## Introduction

Following the spread of the Covid-19 disease in the spring of 2020, most European countries emphasized social distancing among their protective measures. As a consequence, it was not possible to play soccer in those countries. However, studies showed that the risk of spreading the virus during play is minimal (Randers et al. 2021, Gonçalves et al. 2020). Accordingly, both recreational and professional soccer could be played in many countries during the late spring and summer.

In recreational soccer, only a limited number of people could play and train together. Furthermore, social distancing was emphasized off the field, meaning that the use of changing rooms and showering was limited to fewer people.

In professional soccer, protocols were produced to ensure a safe return to play. An example in English has been produced by Department for Digital, Culture, Media & Sport in the United Kingdom (Guidance: Elite sport Stage 1 to 5, 2020). Effort was made to test the players and staff at least once per week. The rationale behind this is that if players are tested frequently, the infection will not enter the group of players and, hence, they do not need to comply with the same social distancing-rules as the rest of the society when being inside their club. However, it is not known what the minimal testing frequency should be, to avoid virus entering the group.

Accordingly, the purpose of the present study is to investigate the incidence of Covid-19 infections in professional soccer players and to compare this to the incidence of the normal population.

## Methods

The top leagues from five different countries where included in the study; England, Sweden, Denmark, Germany and Russia. The countries were selected based on the availability of data on the number of infected players or infected players & staff during the 2020/21-season. The period analyzed was from the beginning of the season until 3rd of January 2021. For the Russian League, data from april through September was used.

Data on the normal population, i.e. number of infected or dead per week was retrieved from the homepage of the European Centre for Disease Prevention and Control (ECDC) (https:\\opendata.ecdc.europa.eu/covid19/nationalcasedeath/xlsx)

Incidence for the normal population was estimated, based on the number of deaths, given an infection-fatality rate of 0.35% (Streeck et al. 2020).

The incidence rate for football players was calculated as the number of positive tests divided by the total number of players in a league. Data was found from different sources; League-homepages, Club-homepages and media-appearances. In cases where data is only available for a group consisting of both players and staff (English Premier League), the incidence rate for one week was calculated as the number of positive tests divided by the number of tests in one week. In cases where players and staff are tested more than once per week, the number of tests is higher than people in that group, leading to an under-estimation of the incidence.

The relative risk was calculated, based on the incidence (or estimated incidence) with 95% confidence intervals.

## Results

The incidence for soccer players was between 9.1% and 14.5% and the estimated incidence for the normal population was between 3.7% and 14.3%.

The relative risk, based on the estimated number of infected people varied between 0.9 and 3.49. A relative risk significantly above 1 was found in Denmark, Sweden and Russia. Table 1 shows a summary of the results from each country.

**Table 1.** Summary of the results from each country. An asterisk (*) shows a relative risk that is significantly greater than 1.

| Country | Incidence, Soccer players [%] | Estimated incidence, normal population [%] | Relative risk (estimate) | Confidence interval (estimate) |
| --- | --- | --- | --- | --- |
| Denmark | 9.9 | 3.7 | 2.56 * | [1.80 - 3.62] |
| Germany | 9.1 | 8.7 | 1.05 | [0.78 - 1.41] |
| England | 10.4 | 14.3 | 0.90 | [0.79 - 1.02] |
| Russia | 14.5 | 3.8 | 3.49 * | [2.91 - 4.18] |
| Sweden | 13.4 | 7.2 | 1.77 * | [1.50 - 2.08] |

## Discussion

Our calculations show that the estimated relative risk of being infected with the Covid-19 virus is larger in soccer players that in the normal population in Denmark, Sweden and Russia.

Estimating the true number of infected people in a population can be done in several ways. We chose to estimate on the basis of deaths, since this seems to be the most consistent number. Other estimations include the percentage of positive tests or blood samples in groups of subjects. These estimations seem to give varying results. However, when estimating the incidence in the normal population, it should be considered, that the reporting of covid-deaths is potentially not the same between countries.

Soccer players are tested frequently, and this should prevent the virus from entering a group of players. However, this does not seem to be the case. There are two possible explanations; either, the tests should be conducted more frequently, or the virus can spread before it can be detected in a test.

In the Guidance for Elite Sport in the United Kingdom, it is emphasized that “all individuals must abide by government and PHE guidelines whilst away from the Competition Venue”. By doing so, soccer players will minimize the risk of being infected, when away from the club. As seen in the results, both soccer players and the normal population get infected. It can be discussed whether or not soccer players should comply better to the recommendations.

## Data Availability

Data is available as stated in the manuscript.

